# In-country cerebrospinal fluid metagenomic next generation sequencing for pathogen detection in Lima, Peru

**DOI:** 10.64898/2026.11.13.26367457

**Authors:** Sumanth P. Reddy, Antonio Chavez Neyra, Katherine Lugo, Brenda Ayzanoa, Carolina Cucho Espinoza, Jarod Morgenroth-Rebin, Oliver A. Bocanegra, Diego Cuicapuza Arteaga, Marlon Saavedra, Chloe Gerungan, Kelsey C. Zorn, Felicia C. Chow, Felix Llanos Tejada, Andres G. Lescano, Pablo Tsukayama, Meddly L. Santolalla, Michael R. Wilson, Renzo Villanueva-Villegas

## Abstract

**Background:** The epidemiology of central nervous system (CNS) infections in low- and middle-income countries is poorly characterized due to limited access to specialized neurodiagnostic testing. Cerebrospinal fluid (CSF) metagenomic next-generation sequencing (mNGS) can detect any pathogen in a single hypothesis-free assay but has largely been confined to high-income countries due to cost and infrastructure requirements. We piloted an in-country CSF mNGS protocol in Lima, Peru to assess the feasibility of this approach in a resource-limited setting.

**Methods:** Adults with suspected CNS infection at Hospital Nacional Dos de Mayo were enrolled. CSF mNGS (DNA and RNA) was performed in-country on a benchtop sequencer with half-volume library reactions and cloud-based analysis (CZID). Paired sequencing at a significantly higher depth was performed on a subset of samples in the United States for validation.

**Results:** Among 43 participants (median age 48 years; 21% living with HIV), in-country mNGS detected 20 pathogens in 18 samples (42%), of which 65% were not identified by routine clinical testing. Nine of 15 patients treated empirically for tuberculous meningitis were found to have a non-mycobacterial infection. Detected organisms included *Toxoplasma gondii*, *Taenia solium*, *Treponema pallidum*, herpesviruses, and, on deeper sequencing, Venezuelan equine encephalitis virus. Deep sequencing recovered four pathogens missed at shallower-depth.

**Conclusions:** In-country CSF mNGS was feasible in Lima, Peru and recovered a clinically and epidemiologically meaningful spectrum of CNS pathogens, most missed by routine testing. By utilizing available, lower-output sequencers, we overcame a principal barrier to deploying this technology where the burden of undiagnosed CNS infection is greatest.

## BACKGROUND

Central nervous system (CNS) infections are commonly encountered in clinical practice in low- and middle-income countries (LMICs) such as Peru. The epidemiology of these illnesses is poorly characterized due to the wide range of endemic pathogens, a significant immunosuppressed population that is at risk for unusual and opportunistic infections, and limited access to specialized neurodiagnostic testing. HIV-associated opportunistic infectious diseases such as tuberculous meningitis (TBM), cerebral toxoplasmosis, and cryptococcal meningitis account for a significant portion of cases, in addition to neurocysticercosis, neonatal bacterial meningitis, and enteroviral and arboviral meningoencephalitis.^1–6^ The poor sensitivity of frontline diagnostic tests for TBM, such as cerebrospinal fluid (CSF) GeneXpert, pose a particular clinical challenge, given the widespread prevalence of TB in urban Peru.^7–9^ Since molecular and serologic CSF testing is limited, patients with subacute meningitis who test negative on CSF GeneXpert are often still thought to have TBM and empirically treated as such. A subset of these patients have alternate non-mycobacterial CNS infections, often leading to suboptimal outcomes.^10^ Improved pathogen characterization could help guide appropriate treatment, inform targeted diagnostic testing algorithms, and contribute to the epidemiological data available in Peru.

CSF metagenomic next generation sequencing (mNGS) is an increasingly important diagnostic tool in high-income settings, and several studies have demonstrated the utility of mNGS as a research-based tool to better understand the epidemiology of CNS infections in LMICs.^11,12^ By broadly amplifying and sequencing the nucleic acid present in a CSF sample, mNGS can theoretically identify any pathogen present, including unexpected and divergent organisms, without requiring an *a priori* hypothesis about the causative pathogen. In Peru, for example, a recent study of CSF mNGS identified pathogens in nearly half of participants with HIV-associated neurologic disease.^5^ Similar studies in Bangladesh, Uganda, Vietnam, and other LMICs have further demonstrated how CSF mNGS is uniquely able to provide valuable epidemiologic information, often uncovering unexpected findings.^11,13–16^ Notably, however, these studies rely on sequencing performed at specialized laboratories in high-income countries rather than locally in the countries where the patients are treated and the samples are collected.

The challenges with performing mNGS locally in LMICs are numerous: sequencing equipment and required consumables are often cost prohibitive, laboratory facilities are in short supply, and expertise and training opportunities for researchers are limited.^11^ Identifying solutions to increase the accessibility of mNGS would advance our understanding of the global burden of neuroinfectious diseases. This study sought to pilot an in-country, research-based CSF mNGS protocol in Lima, Peru, on a lower cost benchtop sequencer as a proof-of-concept to evaluate whether this technology is feasible in a resource limited setting.

## METHODS

### Study population

Participants were enrolled at Hospital Nacional Dos de Mayo (HNDM), a large public hospital in central Lima, Peru, and the study protocol was approved by the institutional review boards of the University of California San Francisco (#24-41322), HNDM (Registro #27060, Código #8967), and Universidad Peruana Cayetano Heredia (UPCH, #197-17-25). It is estimated that 300-400 patients are admitted annually with a suspected CNS infection, but fewer than 25% of patients receive a microbiologically confirmed diagnosis. The hospital’s available diagnostic testing for neurologic infections included Gram stain, bacterial/fungal/AFB culture, India ink staining, GeneXpert MTB/RIF (Cepheid), and FilmArray PCR (BioFire Diagnostics, includes *Escherichia coli K1, Haemophilus influenzae, Listeria monocytogenes, Neisseria meningitidis, Streptococcus agalactiae, Streptococcus pneumoniae,* cytomegalovirus (CMV), herpes simplex virus-1 (HSV-1), herpes simplex virus 2 (HSV-2), human herpesvirus 6 (HHV-6), enterovirus, human parechovirus, varicella zoster virus (VZV), and *Cryptococcus neoformans/gattii*). Admitted adults who underwent lumbar puncture due to suspected CNS infection from August 2024 - April 2025 were screened for participation. Suspected CNS infection was determined by the treating clinician and defined pragmatically as any adult undergoing lumbar puncture for evaluation of subacute or acute neurologic symptoms (e.g., headache, fever, meningismus, altered mental status, seizures, or focal neurologic deficits) in whom a CNS infection was in the differential diagnosis. After participants or their family members provided informed consent, approximately 0.5 – 2 mL of leftover CSF was collected and stored at -80°C after all ordered clinical testing was in process. Participants were excluded if less than 500µL of surplus CSF was available, CSF was stored at 4°C for greater than 48 hours before transfer to -80°C, medical records were unavailable for review, or antibiotics were administered for 7 or more days prior to lumbar puncture. Possible, probable, and definite TBM diagnoses were defined by the Uniform Case Definition.^17^

DNA/RNA extraction and library preparation was performed in batches at UPCH (a local university located approximately 7 km from HNDM) according to a previously published protocol (**Figure 1**).^18,19^ To minimize reagent costs, the library preparation reactions were performed at half-volumes. The library concentrations were measured via Qubit fluorometer, and a small aliquot of the library was over-amplified to allow for visualization of library size via agarose gel electrophoresis. The DNA libraries were pooled and sequenced on two NextSeq 550 (Illumina) 300-cycle paired-end high-output sequencing runs, whereas all RNA libraries were pooled and sequenced on a single run with a goal of approximately 20-30M and 10-15M paired-end reads for each DNA and RNA library, respectively. Sequencing data were processed via the open-source, cloud-based metagenomics platform, CZID,^20^ and pathogen calls were determined via a previously published methodology.^18^

**Figure 1:**
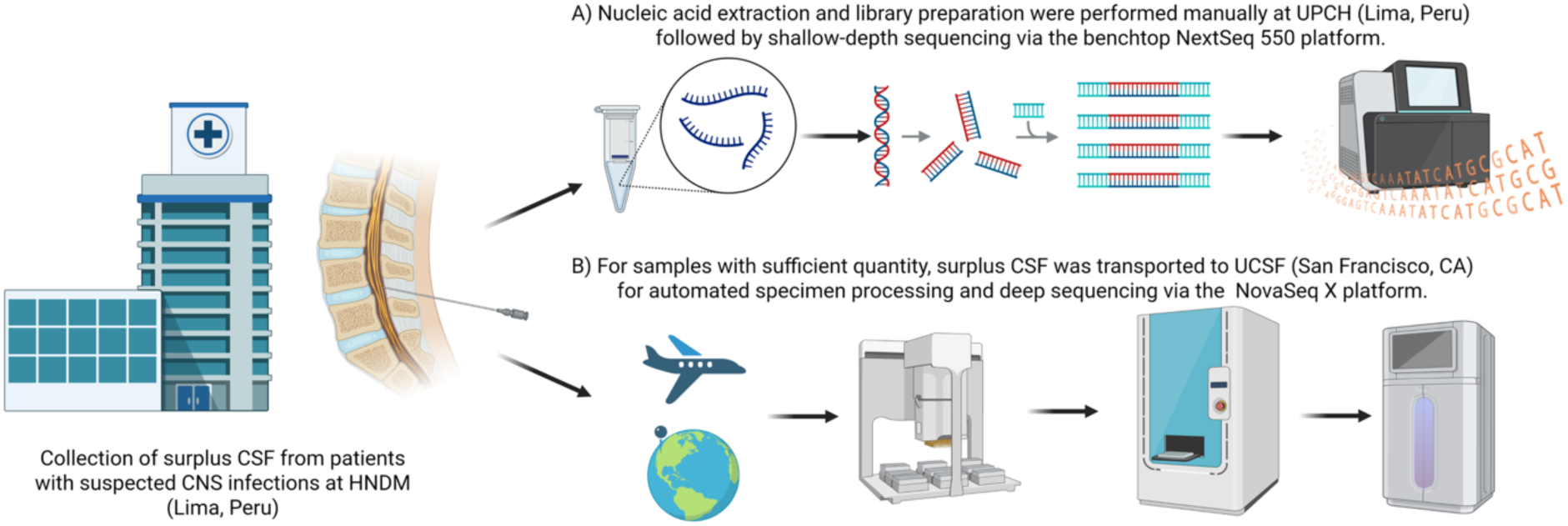
Study design and sequencing approaches

A subset of participants had more than one aliquot of surplus CSF. These aliquots were stored in a nucleic acid preservative and pathogen inactivating reagent, DNA/RNA Shield (Zymo, Cat# R1100-50), and transported to the United States at approximately - 20°C. After extraction and library preparation using full reagent volumes according to the manufacturer’s protocol, the RNA and DNA libraries were deep sequenced on two separate lanes of a 300-cycle NovaSeq X high-output sequencing run with a goal of approximately 150M paired-end reads for each DNA and RNA library. The data were processed and analyzed in the same manner as described above.

## RESULTS

### Cohort overview

Of the 43 participants included in the study (**Figure 2**), 11 were female (26%) and the median age was 48 years (IQR: 37-57, **Table 1**). There were 9 participants (21%) living with HIV with a median CD4 count of 24 cells/mm^3^ (range: 17-979) and a median viral load of 13,300 copies/mL (range: undetectable – 157,000). Six participants living with HIV were diagnosed during this same admission. Most patients (n=36, 84%) had no neurologic symptoms or no significant disability (mRS 0-1) prior to the onset of current symptoms. The most common presenting symptoms were headache (n=39, 91%), altered mental status (n=33, 77%), and neck stiffness (n=22, 51%). CSF analysis commonly demonstrated mild pleocytosis (median=10 cells/mm^3^, IQR: 3-88 cells/mm^3^), normal glucose (median=49 mg/dL, IQR: 40-58 mg/dL), and elevated protein (median=116 mg/dL, IQR 88-292 mg/dL).

**Figure 2:**
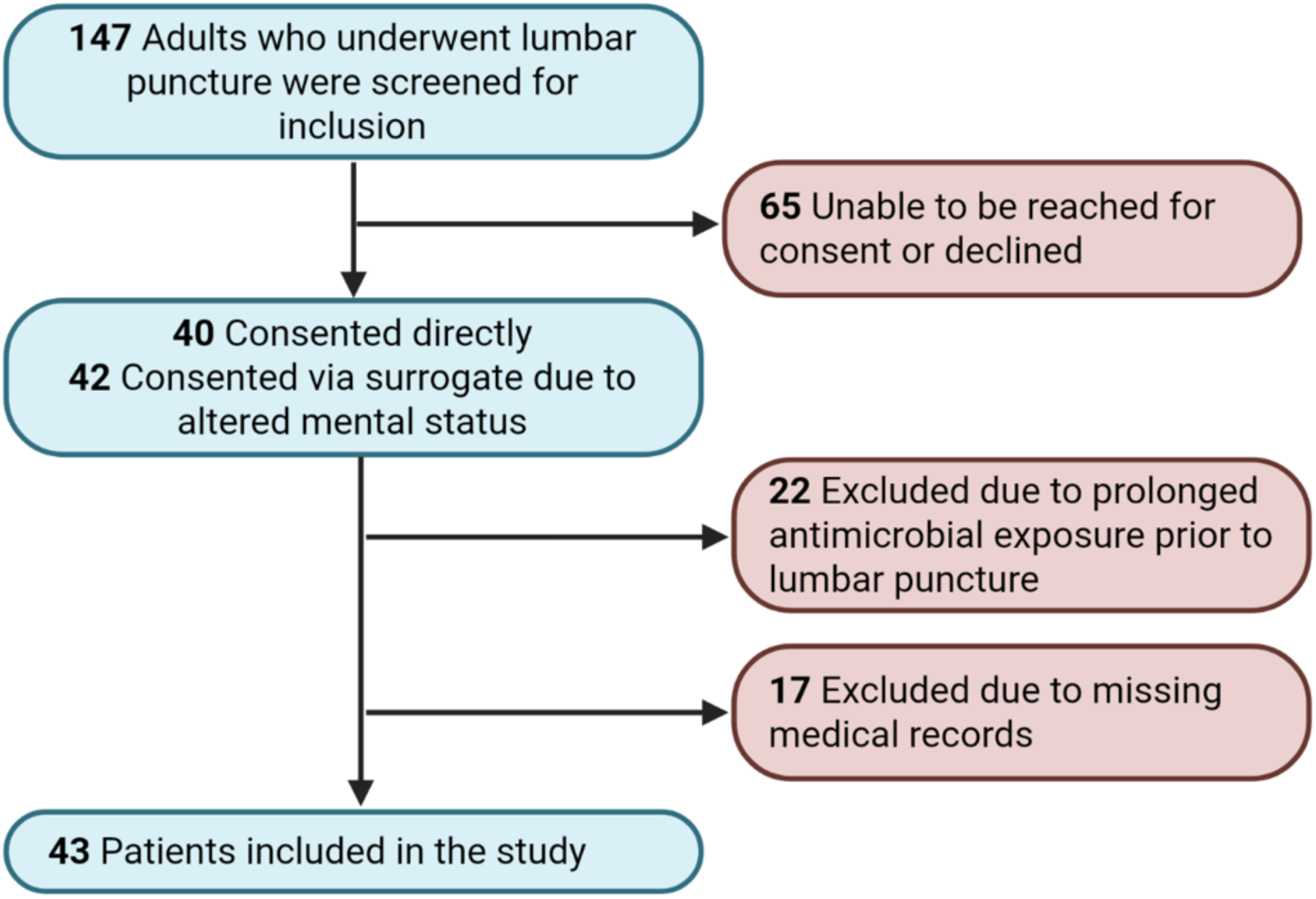
Participant flow diagram

**Table 1:** Baseline demographic and clinical characteristics.

| **Characteristic** | **Overall (N=43)** | **CNS infection identified via mNGS or clinical testing (n=23)** | **No infection identified (n=20)** |
|---|---|---|---|
| Age, years (n=43) | 48.5 ± 15.7; median 48.0 [IQR 37.0–57.0] | 49.5 ± 15.6; median 48.0 [IQR 40.5–56.5] | 47.4 ± 16.3; median 49.5 [IQR 34.2–57.2] |
| Male | 32 (74.4%) | 19 (82.6%) | 13 (65.0%) |
| Female | 11 (25.6%) | 4 (17.4%) | 7 (35.0%) |
| **Residence** |  |  |  |
| Lima | 37 (86.0%) | 20 (87.0%) | 17 (85.0%) |
| Outside Lima | 6 (14.0%) | 3 (13.0%) | 3 (15.0%) |
| **Prior medical history — no. (%)** |  |  |  |
| Hypertension | 6 (14.0%) | 4 (17.4%) | 2 (10.0%) |
| Diabetes | 3 (7.0%) | 1 (4.3%) | 2 (10.0%) |
| Stroke | 2 (4.7%) | 1 (4.3%) | 1 (5.0%) |
| CAD/MI | 0 (0.0%) | 0 (0.0%) | 0 (0.0%) |
| Tuberculosis | 6 (14.0%) | 2 (8.7%) | 4 (20.0%) |
| **HIV status (N=43)** |  |  |  |
| Positive (new diagnosis) | 6 (14.0%) | 4 (17.4%) | 2 (10.0%) |
| Positive (known prior to admission) | 3 (7.0%) | 3 (13.0%) | 0 (0.0%) |
| Negative | 34 (79.1%) | 16 (69.6%) | 18 (90.0%) |
| **Symptoms at presentation** |  |  |  |
| Headache | 39 (90.7%) | 22 (95.7%) | 17 (85.0%) |
| Neck stiffness | 22 (51.2%) | 15 (65.2%) | 7 (35.0%) |
| Photophobia | 2 (4.7%) | 0 (0.0%) | 2 (10.0%) |
| Any altered mental status | 33 (76.7%) | 18 (78.3%) | 15 (75.0%) |
| Coma | 0 (0.0%) | 0 (0.0%) | 0 (0.0%) |
| Cranial nerve palsy | 6 (14.0%) | 2 (8.7%) | 4 (20.0%) |
| Hemiparesis/hemiplegia | 7 (16.3%) | 5 (21.7%) | 2 (10.0%) |
| Sensory loss | 4 (9.3%) | 2 (8.7%) | 2 (10.0%) |
| Ataxia/dysmetria | 2 (4.7%) | 1 (4.3%) | 1 (5.0%) |
| **Laboratory Results, median [IQR]** |  |  |  |
| Serum WBC, cells/μL | 9033.0 [6801.0–11982.0] | 8813.0 [6893.0–11146.0] | 9095.5 [6295.2–12815.2] |
| CSF WBC, cells/μL | 10.0 [3.0–87.5] | 20.0 [2.5–67.5] | 9.0 [3.0–178.5] |
| CSF glucose, mg/dL | 49.0 [39.5–57.5] | 45.0 [39.5–53.5] | 55.5 [40.2–63.0] |
| CSF protein, mg/dL | 116.0 [87.8–291.5] | 129.0 [79.8–291.5] | 112.0 [90.5–251.2] |

The median length of hospital admission was 25 days (IQR 13-54.5), and median duration of follow up from the time of admission was 148 days (IQR: 41-383). Three participants (7%) died during the hospitalization, and 8 (18.6%) had died during the follow up period. Eleven participants required intubation (25.6%), and 8 (18.6%) required CSF diversion via either external ventricular drainage or ventriculoperitoneal shunting. Nearly all participants experienced a worsening in their functional status at the time of discharge compared to their premorbid baseline (**Figure 3D**).

**Figure 3:**
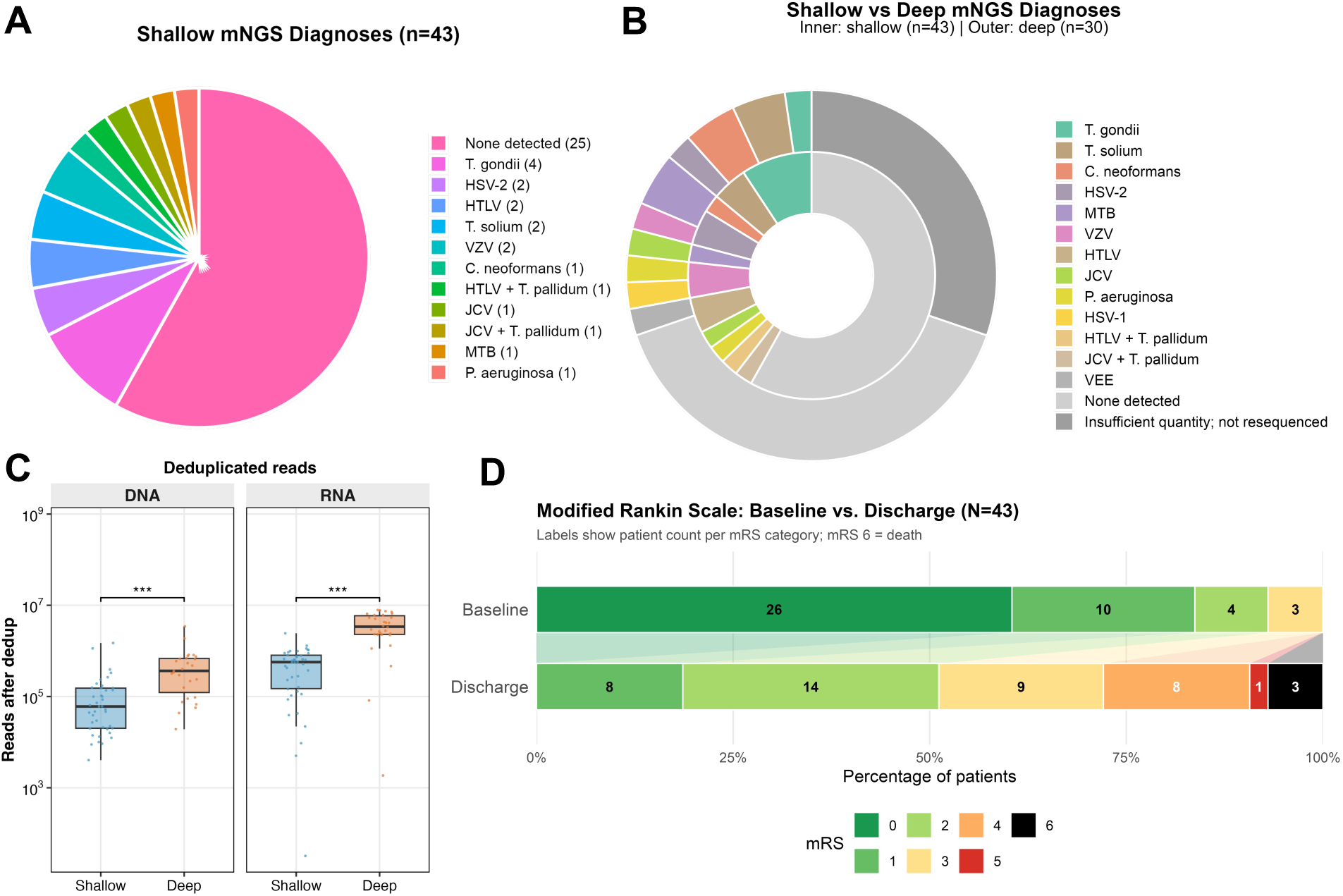
A) Pathogens detected via shallow-depth mNGS, B) compared to deep sequencing. C) The number of reads was substantially higher with deep sequencing, and this remained the case when accounting for duplicate reads. D) Patient outcomes prior to their neurologic illness, compared to at discharge, via modified Rankin Scale (mRS)

### mNGS results

CSF mNGS was performed in Peru to a median depth of 28M DNA reads (IQR: 18M - 38M) and 12M RNA reads (IQR: 10M - 13M). A total of 20 pathogens were identified in 18 (42%) of samples, including *Toxoplasma gondii* (n=4), HSV-2 (n=2), human T-lymphotropic virus 1 (HTLV-1) (n=3), *Taenia solium* (n=2), and VZV (n=2), among others (**Figure 3A**). Two patients had multiple pathogens detected via mNGS (HTLV-1 and *Treponema pallidum*; JC virus and *T. pallidum*). Among pathogens detected via shallow-depth mNGS, 13 (65%) were not detected on routine clinical testing during the hospital admission. Within the limitations of the available clinical data, the positive detections correlated with the clinical presentation with the exception of two cases of HTLV-1, which were felt to be incidental detections in the setting of blood contamination rather than the cause of the neurologic presentation (**Supplemental Appendix**). Nine out of 15 patients with possible/probable TBM^17^ who were treated with empiric TBM therapy were found to have a non-mycobacterial infection via mNGS. Shallow-depth mNGS confirmed the only TBM diagnosed via GeneXpert MTB/RIF, but missed a case of known cryptococcal meningitis and failed to identify the cause of an intracranial abscess in a patient with multiple myeloma, which was later diagnosed as *Listeria monocytogenes* via culture from surgical drainage (**Supplemental Appendix**).

Surplus CSF was available for 30 patients for deep sequencing at UCSF, which was performed to a median depth of 113M DNA reads (IQR: 39M - 150M) and 151M RNA reads (IQR: 75M - 155M). After quality filtering and de-duplication, the average number of DNA and RNA reads was 3.7 and 6.9 times greater, respectively with deep sequencing compared to shallow-depth sequencing (p < 0.001, **Figure 3C**). Pathogens were detected in 47% of samples with deep sequencing, including 4 that were missed on shallow-depth sequencing: *Cryptococcus neoformans*, *M. tuberculosis*, HSV-1, and an uncommon pathogen, the mosquito-borne alphavirus, Venezuelan equine encephalitis virus (VEEV). Two pathogens that were detected on shallow-depth sequencing were missed on deep sequencing, *T. pallidum* and *T. gondii*.

### Selected cases

#### Venezuelan equine encephalitis virus

A man aged 71-75 years with angiosarcoma of the liver who had undergone surgical resection six months prior presented with three days of fever and one day of disorientation and word-finding difficulty after recently returning from the Peruvian Amazon. CSF was notable only for an elevated protein of 115 mg/dL, with a normal cell count and glucose. Brain magnetic resonance imaging (MRI) was not obtained. The working diagnosis was metabolic encephalopathy, given his history of liver cancer, and the lactulose dose was increased. He was discharged 7 days later with minimal change in his mental status. While no pathogens were identified on shallow-depth sequencing, deep sequencing identified 794 RNA reads mapping to one region of the VEEV genome, spanning a contiguous region of approximately 190 base pairs within the nsP1–nsP2 replicase junction (genome positions 1930–2120); BLASTN analysis confirmed that all matching reads aligned exclusively to VEEV reference sequences, with no significant similarity to other alphavirus species.

### T. gondii

A man aged 51-55 years with a recent diagnosis of pulmonary TB, on treatment, presented with three months of cough, weight loss, and constitutional symptoms, one month of headache and neck stiffness, and one day of being unable to answer basic questions. He was newly diagnosed with HIV on admission (viral load and CD4 count unknown). CSF showed 5 WBC/µL, a protein of 228 mg/dL, and a glucose of 39 mg/dL. Neuroimaging revealed an isolated ring-enhancing lesion in the right cerebellar hemisphere (**Figure 4a**). He was treated as probable TB meningitis and was lost to follow-up after he was discharged 52 days later with minimal change in his symptoms. Shallow-depth sequencing detected *T. gondii*, which uncommonly presents as an isolated lesion in the infratentorium (surplus CSF was not available for confirmatory deep sequencing).

**Figure 4:**
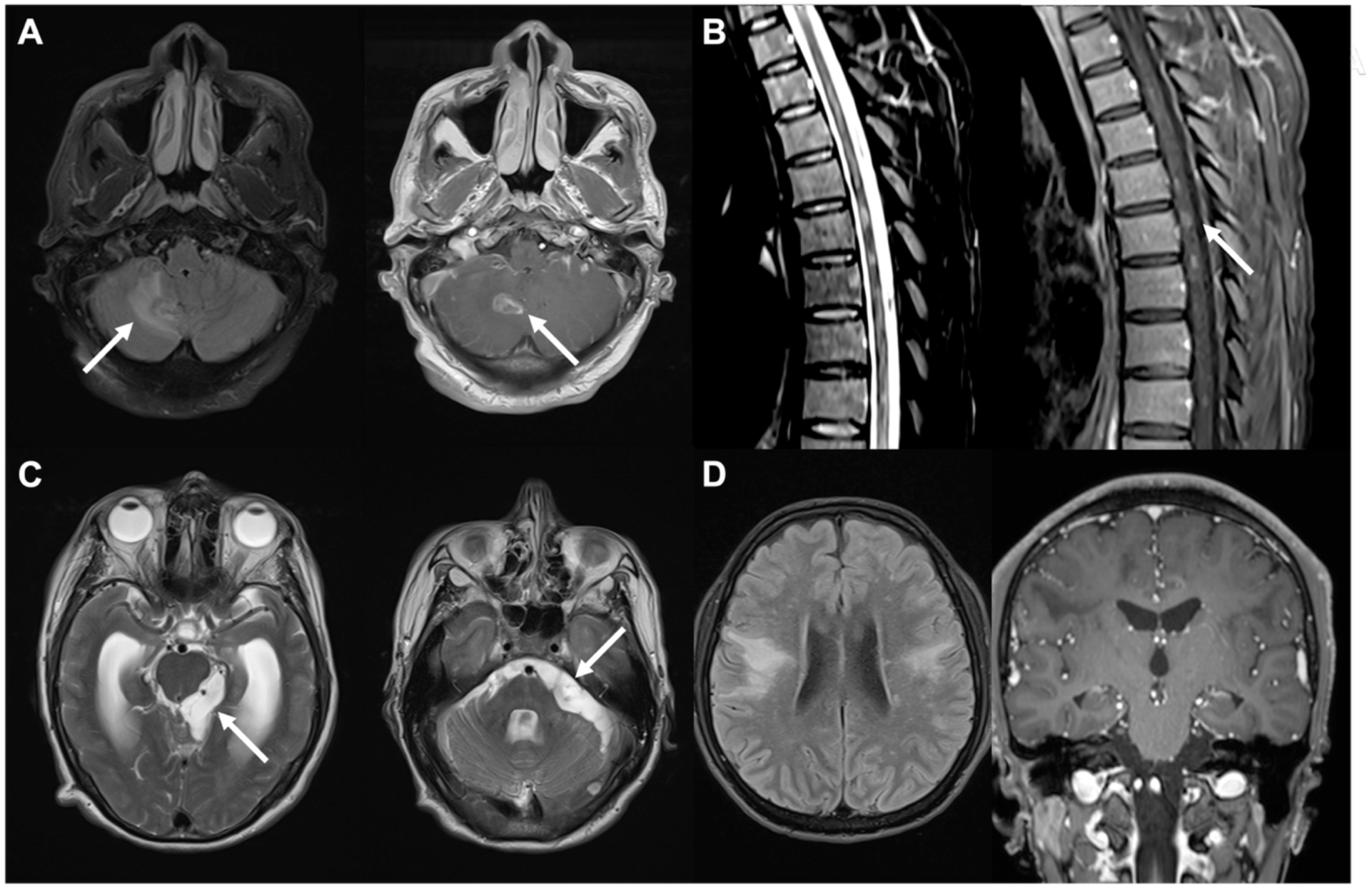
Magnetic resonance imaging (MRI) from select cases. A) T2/FLAIR (right) and T1 post-contrast imaging (left) demonstrating a T2 hyperintense cerebellar ring enhancing lesion from a patient ultimately found to have cerebral toxoplasmosis on mNGS. B) T2/FLAIR (right) and T1 post-contrast imaging (left) indicating T2 hyperintense lesions with patchy contrast enhancement throughout the thoracic cord. The patient was ultimately found to have *Treponema pallidum* and HTLV-I on CSF mNGS. C) T2 images demonstrating cyst-like structures in the subarachnoid space surrounding the brainstem and cerebellum, consistent with a diagnosis of racemose neurocysticercosis. D) T2/FLAIR (right) and T1 post-contrast imaging (left) demonstrating bi-hemispheric non-enhancing lesions. This patient was living with advanced HIV (CD4 count 17 cells/mm^3^) and passed away 3 months after presentation despite prolonged treatment with broad spectrum antimicrobials. Shallow-depth CSF mNGS ultimately detected JC virus, consistent with a diagnosis of progressive multifocal leukoencephalopathy (PML).

### *T. pallidum* with HTLV-1 co-detection

A man aged 31-35 years presented with several weeks of headache and lower-extremity weakness and sensory loss. CSF showed 2 WBC/µL and a protein of 129 mg/dL. Spinal MRI demonstrated foci of hyperintense T2 signal within the thoracic cord with post-contrast enhancement (**Figure 4b**). He was treated with plasmapheresis and corticosteroids without improvement and discharged wheelchair-dependent 30 days later. Subsequent shallow-depth sequencing detected both *T. pallidum* and HTLV-1; both pathogens were confirmed on deep sequencing as well. Syphilitic myelitis is a rare manifestation of neurosyphilis (on the order of ∼1.5% of cases) and characteristically produces longitudinally extensive T2 signal with superficial, peripheral cord enhancement, the pattern seen here. HTLV-1– associated myelopathy/tropical spastic paraparesis (HAM/TSP), by contrast, is generally (but not always) a chronic progressive myelopathy whose hallmark is cord *atrophy* rather than T2 signal hyperintensity or enhancement. Therefore, the acute, swollen, and enhancing long-segment lesion was felt to be most likely due to the syphilitic process, with HTLV-1 regarded as a concurrent (and possibly chronic and contributory) infection rather than the driver of the acute presentation.

### T. solium

A man aged 51-55 years without significant past medical history presented with two weeks of nausea, vomiting, headache, and one day of somnolence requiring noxious stimulation to arouse. On presentation, testing demonstrated a mild peripheral leukocytosis (11.5 × 10⁹/L) and CSF with 30 WBC/µL (6% neutrophils, 94% mononuclear cells), a protein of 269 mg/dL, and a glucose of 24 mg/dL. MRI demonstrated hydrocephalus with basal meningeal enhancement with cyst-like structures, most suggestive of racemose neurocysticercosis (**Figure 4c**). He was discharged on albendazole and high-dose prednisone. Shallow-depth sequencing detected *T. solium* with numerous aligned reads, confirming the diagnosis.

## Discussion

In this study we show that CSF mNGS can be performed locally to reveal a clinically and epidemiologically meaningful spectrum of CNS pathogens in Lima, Peru. Across 43 samples, mNGS performed locally detected at least one plausible pathogen in 42% of samples, and the majority of these organisms had not been identified by routine clinical testing performed during the admission. The detections spanned DNA and RNA viruses, *Mycobacterium tuberculosis*, *C. neoformans*, and organisms that are difficult to diagnose with conventional assays: *T. pallidum*, *Taenia solium*, and *Toxoplasma gondii*. CSF mNGS is established as a diagnostic capable of detecting a broad range of pathogens, and a recent single-center study demonstrated that approximately 20% of patients with CNS infections at a U.S. tertiary medical center were diagnosed via mNGS alone, underscoring the importance of this approach even in resource-rich settings.^21–23^ Our findings extend that premise in two directions that matter for the settings where the diagnostic need is greatest: the sequencing was performed locally rather than sent to a reference center abroad, and it was performed at a depth low enough to allow for sequencing to be batched and performed on less costly and available equipment.

Samples were sequenced to a median of approximately 28 million DNA and 12 million RNA reads, roughly 3.7-fold and 6.9-fold fewer de-duplicated reads than the paired deep-sequencing runs performed on the validation subset. mNGS at a sequencing depth that is more practical in Peru allows for the assay to be run on low-output benchtop sequencers such as the iSeq, MiSeq, or NextSeq at an order-of-magnitude lower acquisition cost, a substantially smaller instrument footprint, and far lower service, maintenance, and reagent-commitment burdens than high-output platforms such as NovaSeq and similar platforms. The capital expense and the ongoing service contracts of high-output sequencers are a primary reason clinical mNGS has remained concentrated in well-resourced reference laboratories.

This accessibility comes at a cost of sensitivity – on the paired validation subset, deeper sequencing recovered four pathogens that shallower sequencing did not (*C. neoformans*, *M. tuberculosis*, HSV-1 and VEEV). The organisms missed at shallower depth were, as expected, those present at low genomic abundance in CSF, and laboratory validation and large prospective series of CSF mNGS have similarly shown that detection at the limit of the assay is governed by pathogen read abundance and input quality.^22,24^ Therefore, shallower-depth mNGS is best understood not as a replacement for deeper sequencing where maximal sensitivity is optimized, but as a high-throughput, lower-cost screening modality that trades low-end sensitivity for the accessibility that allows the assay to be operated locally.

An important limitation of CSF mNGS is highlighted by two detections, *T. pallidum* and *T. gondii* (both in participants distinct from the cases above), which were made on shallower-depth sequencing but not recovered on subsequent deeper sequencing of separate CSF aliquots. Rather than indicating false-positive calls, this discordance likely represents the stochasticity of detecting low-abundance targets from limited nucleic-acid input, compounded by aliquot-to-aliquot variation. This observation tempers the expectation that greater depth uniformly improves sensitivity, and underscores pre-analytic factors such as input volume, aliquoting, and low pathogen burden as primary determinants of detection at the low end.^25^

Prior applications of CSF mNGS in low- and middle-income settings, including the characterization of neurological infection among people living with HIV in Peru, have reported that unbiased sequencing identifies a pathogen in a substantial fraction, approaching half of otherwise-undiagnosed cases.^5^ Our positive-detection rate (42%) is concordant with that experience, and the ability to mirror this yield with a lower-resource workflow strengthens the validity of in-country CSF mNGS.

The clinical relevance of these detections is best illustrated by the subset of patients treated empirically for TBM. Of 15 patients who were managed as presumptive TBM, mNGS identified a non-mycobacterial infection in nine. This pattern reflects a recognized diagnostic difficulty – frontline molecular testing for TBM, including Xpert MTB/RIF Ultra on CSF, is insensitive, missing a substantial proportion of true cases,^7,8,26^ so that a negative result cannot exclude TBM, and patients are frequently continued on empiric, monthslong anti-TB treatment in the absence of a confirmed diagnosis.

Applying a uniform case definition for TBM^17^ to such patients still leaves a substantial group^15,18^ whose true etiology is non-mycobacterial in origin, and mNGS could potentially elucidate the true etiology within this subset.

In addition to sequencing at a shallower-depth, several deliberate choices made the workflow feasible and affordable in this setting, and together they constitute a template that could be reproduced in a variety of settings. Library preparation reactions utilized half the standard reagent volumes, reducing per-sample consumable costs. Sequence analysis used a cloud-based pipeline (CZID),^20^ which obviated the need for local high-performance computing infrastructure and specialized bioinformatics staff.

Several limitations should be emphasized. CSF mNGS was performed months after each patient’s admission as a retrospective research assay and thus, it did not inform real-time clinical management, and all statements of clinical impact are therefore necessarily conditional. The results indicate how unbiased sequencing *could* have impacted clinical care had the results been available prospectively, not that it did contribute. Second, the sample size is modest (n=43), and the cohort was assembled from residual CSF among consenting participants, introducing selection effects that limit epidemiologic generalization. Third, CSF was banked and aliquoted before testing, and pre-analytic factors, such as input volume, freeze-thaw, and low pathogen burden, are expected to contribute to lower sensitivity, as reflected in the shallower-versus-deeper discordance discussed above.^25^ Fourth, orthogonal confirmation was incomplete for some detections, and mNGS identification of one organism may not exclude concurrent infection.

Establishing actual clinical impact ultimately requires prospective, real-time deployment with formal measurement of turnaround time and effects on clinical decision-making. We note, however, that full clinical validation of an mNGS assay is a substantial undertaking: rigorous determination of analytic sensitivity and specificity, limits of detection across organism classes, reproducibility, and the regulatory and quality-system requirements for a laboratory-developed test carry a cost and complexity that are themselves a major barrier.^22^ A pragmatic near-term path is therefore to use mNGS as a research and epidemiologic surveillance tool – for example, in this study, the prevalence of *T. gondii* has raised interest in making a singleplex *T. gondii* PCR clinically available at HNDM. The tremendous diversity of pathogens uncovered in this relatively limited subset of participants further justifies investment in this area to truly understand the global burden of neuroinfectious diseases.

## Disclosures

M.R.W has received research-unrelated grant funding from Roche/Genentech, Novartis, and Kyverna Therapeutics, consulting fees from Ouro Medicines, Indapta Therapeutics, Vertex Pharmaceuticals, Red Tree Ventures, and Pfizer, and is a co-founder and on the Board of Directors for Delve Bio, Inc.

## Funding

This project was supported by the Fogarty International Center of the National Institutes of Health (NIH) under Award Number D43TW009343, the University of California Global Health Institute (UCGHI), the American Academy of Neurology Clinical Research Training Scholarship, and the Westridge Foundation. Sequencing was performed at the UCSF Center for Advanced Technology, supported by UCSF PBBR, RRP IMIA, and NIH 1S10OD028511-01 grants.

## DATA AVAILABILITY

Sequencing data will be available via the Sequence Read Archive (SRA) at the time of publication.

## Supplementary Appendix 1: Clinical Summaries of CSF mNGS Detections

### Patient 001: *Toxoplasma gondii*

A man aged 41-45 with no prior medical history presented with one month of cough, dyspnea, weight loss, intermittent fevers, and night sweats. He also had several weeks of headache and neck stiffness, followed by two days of altered mental status. HIV was diagnosed on admission, with a CD4 count of 60 cells/µL. CSF was acellular, with protein 428 mg/dL and glucose 39 mg/dL. MRI findings were consistent with cerebral toxoplasmosis with multifocal T2/FLAIR lesions with variable peripheral enhancement. T. gondii was detected via shallow and deep mNGS, and HIV RNA was also detected.

### Patient 004: *Cryptococcus neoformans*

A man aged 41-45 presented with 2-3 weeks of weight loss, fever, night sweats, nausea, and vomiting, plus one week of headache, neck stiffness, altered mental status, and dysphagia. HIV was newly diagnosed on admission (CD4 count and viral load were not available). CSF showed 245 WBC/µL (100% mononuclear), protein 393 mg/dL, and glucose 37 mg/dL, and CSF culture grew Cryptococcus neoformans. Both shallow and deep mNGS detected C. neoformans and HIV.

### Patient 009: HSV-2

A man aged 21-25 with no prior medical history presented with several days of nausea, headache, and neck stiffness. CSF showed 350 WBC/µL (98% mononuclear, 2% neutrophils), protein 257 mg/dL, and glucose 48 mg/dL, and BioFire FilmArray was positive for HSV-2. mNGS detected HSV-2 via shallow RNA and deep mNGS, matching the clinical diagnosis of HSV-2 meningitis.

### Patient 011: *Treponema pallidum* and JC polyomavirus

A man aged 46-50 with HIV on ART (CD4 979 cells/µL), presented with weeks of headache, neck stiffness, and altered mental status. CSF parameters were unremarkable. No neuroimaging was obtained. Serum RPR was positive (titer unknown), and he was treated empirically for neurosyphilis. mNGS detected T. pallidum via shallow mNGS, supporting the clinical suspicion of neurosyphilis. JC polyomavirus was also detected, though without neuroimaging, its significance is uncertain.

### Patient 014: HTLV-1 and *Treponema pallidum*

Described in detail in the main text.

### Patient 024: HTLV-1

A man aged 76-80 presented with two days of headache, neck stiffness, and altered mental status. CSF showed 65 WBC/µL (100% mononuclear), 50 RBC/µL, protein 472 mg/dL, and glucose 45 mg/dL, with xanthochromia. Further workup revealed subarachnoid hemorrhage from a ruptured intracranial aneurysm. mNGS detected HTLV-1 via shallow mNGS. Because HTLV-1 is endemic in Peru, this was considered most consistent with incidental detection from blood-derived lymphocytes in a probable HTLV-1 carrier, not a cause of the presentation.

### Patient 035: JC polyomavirus

A man aged 26-30 with HIV (CD4 17 cells/µL, viral load 157,000 copies/mL) presented with headache, confusion, and asymmetric upper extremity weakness (left greater than right). CSF parameters were unremarkable. MRI showed bihemispheric non-enhancing lesions (Figure 4D). He died three months after presentation despite prolonged broad-spectrum antimicrobial therapy. Both shallow and deep mNGS detected JC polyomavirus and HIV. Advanced immunosuppression, bland CSF, non-enhancing multifocal white matter lesions, and progression despite antimicrobial therapy are characteristic of progressive multifocal leukoencephalopathy, which supports JCV as the cause.

### Patient 045: HTLV-1

A man aged 51-55 with hypertension presented with one day of headache, neck stiffness, and altered mental status. CSF showed 70 WBC/µL (90% mononuclear, 10% neutrophils), protein 49 mg/dL, and glucose 41 mg/dL. Brain CT reports described possible ventriculitis and hydrocephalus; the images were not available for review. Subarachnoid hemorrhage was suspected, but no aneurysm was found. He was managed conservatively and required an external ventricular drain, followed by ventriculoperitoneal shunting prior to discharge. mNGS detected HTLV-1 via shallow mNGS. As in Patient 024, this was considered most consistent with incidental detection of HTLV-1 from blood in a probable carrier.

### Patient 048: *Mycobacterium tuberculosis*

A man aged 41-45 with no prior medical history presented with several weeks of headache and cognitive change. HIV was newly diagnosed on admission (CD4 count unavailable). CSF parameters were unremarkable. MRI showed diffuse frontoparietal T2/FLAIR signal abnormality and mild basilar meningeal enhancement. He worsened despite empiric vancomycin and meropenem for suspected bacterial meningitis, and later empiric antituberculous therapy, and died about one month after presentation. mNGS detected M. tuberculosis (deep sequencing only) in addition to HIV. Basilar meningeal enhancement and the subacute fatal course in a newly diagnosed HIV patient are consistent with TBM.

### Patient 049: *Cryptococcus neoformans*

A man aged 56-60 with HIV (CD4 24 cells/µL, viral load 48,300 copies/mL) presented with two weeks of fever, chills, and weight loss, plus headache and confusion. CSF profile was notable for protein of 1,097 mg/dL. MRI showed hydrocephalus and nodular subcortical foci in both frontal lobes. Cryptococcal meningitis was confirmed by India ink. Shallow-depth mNGS ***did not*** detect C. neoformans; it was detected only via deep sequencing. Since the diagnosis was microbiologically confirmed, this deep-only detection represents true recovery of a pathogen missed at shallow depth.

### Patient 069: Venezuelan equine encephalitis virus

Described in detail in the main text.

### Patient 070: HSV-2

A man aged 26-30 with no prior medical history presented with one month of cough, weight loss, and fever, and was diagnosed with pulmonary tuberculosis based on chest radiograph and a positive AFB sputum culture. He also had one week of severe headache. CSF parameters were unremarkable, and brain MRI was normal. The headache resolved over two weeks without specific treatment. mNGS detected HSV-2 via shallow mNGS. The self-limited course fits HSV-2 meningitis, though no orthogonal clinical validation was available.

### Patient 072: Varicella-zoster virus

A woman aged 81-85 with no known medical history presented with several days of severe headache and mild confusion. CSF showed 42 WBC/µL (100% mononuclear), protein 73 mg/dL, and glucose 76 mg/dL. Brain MRI was unremarkable, and BioFire FilmArray was not performed. She was treated empirically with acyclovir and discharged on hospital day 15 after completing the course, with her symptoms resolved. mNGS detected VZV via shallow-depth sequencing. Lymphocytic pleocytosis, advanced age as a risk factor for VZV reactivation, and recovery on acyclovir together support VZV meningoencephalitis.

### Patient 077: *Taenia solium*

A man aged 66-70 with suspected neurocysticercosis and a prior ventriculoperitoneal shunt presented with several days of recurrent fever and headache. CSF showed 110 WBC/µL (86% mononuclear, 14% neutrophils), protein 130 mg/dL, and glucose 41 mg/dL. MRI showed progression of suspected racemose neurocysticercosis with prominent hydrocephalus and basal meningeal enhancement. mNGS detected Taenia solium via both shallow and deep sequencing approaches, confirming the suspected diagnosis.

### Patient 084: *Toxoplasma gondii*

A man aged 61-65 with no prior medical history presented with one week of headache, one day of left visual field loss, and a generalized tonic-clonic seizure. CSF showed 210 WBC/µL (100% mononuclear), protein 973 mg/dL, and glucose 37 mg/dL. MRI showed multifocal hemispheric T2/FLAIR lesions with variable enhancement. HIV status was not documented, and he left at his own request several days after presentation, before workup was complete. mNGS detected Toxoplasma gondii via shallow-depth sequencing only (missed on deep sequencing). Multifocal enhancing lesions are compatible with toxoplasmosis, but without documented immunosuppression or confirmatory testing, this detection is considered possible.

### Patient 093: *Toxoplasma gondii*

Described in detail in the main text.

### Patient 095: HSV-1

A man aged 36-40 presented with one to two weeks of back pain and one day of headache and neck stiffness. CSF showed 40 WBC/µL (100% mononuclear), protein 314 mg/dL, and glucose 40 mg/dL. Head CT was unremarkable; brain MRI was not obtained. He was treated empirically with vancomycin, meropenem, and acyclovir after lumbar puncture. Spine MRI, obtained about 10 days after admission, showed lumbosacral epidural abscesses with osteomyelitis; abscess culture grew Staphylococcus aureus. Acyclovir was then stopped after nearly a full treatment course. mNGS detected HSV-1 only on deep sequencing. The parameningeal infection likely accounts for much of the CSF abnormality, but a concurrent HSV-1 meningitis, partially or fully treated by the acyclovir he received, cannot be excluded.

### Patient 101: *Toxoplasma gondii*

A man aged 56-60 with recently diagnosed dengue after travel to the Peruvian Amazon presented with two weeks of fever and weight loss, followed by one week of headache and neck stiffness. CSF protein was 139 mg/dL. MRI showed a single peripherally enhancing T2/FLAIR lesion in the left frontal lobe. Because he appeared well and had recently had dengue, the treating team suspected post-infectious encephalitis. HIV status was unknown. He had a persistent headache at one-month clinic follow-up and was then lost to follow-up. mNGS detected Toxoplasma gondii via shallow sequencing. A peripherally enhancing lesion is compatible with toxoplasmosis, and atypical Amazonian T. gondii strains are known to cause severe disease in immunocompetent adults.

### Patient 104: *Taenia solium*

Described in detail in the main text.

### Patient 107: Varicella-zoster virus

A woman aged 46-50 with hypertension, hyperlipidemia, and end-stage renal disease presented with one week of herpes zoster rash, then several days of confusion and mild right hemiparesis, worse in the leg than the arm. CSF showed 10 WBC/µL (52% mononuclear, 48% neutrophils), protein 87 mg/dL, and glucose 40 mg/dL. Head CT was unremarkable. She was treated empirically with acyclovir and responded well. mNGS detected VZV via both shallow and deep sequencing.

### Patient 112: *Pseudomonas aeruginosa*

A man aged 41-45 presented with one week of fever and headache after recent cerebellar tumor resection. CSF showed 20 WBC/µL (90% mononuclear, 10% neutrophils), protein 100 mg/dL, and glucose 49 mg/dL, and CSF culture grew Pseudomonas aeruginosa. Shallow and deep mNGS detected the same organism, supporting a diagnosis of postoperative Pseudomonas meningitis.

### Patient 114: *Mycobacterium tuberculosis*

An immunocompetent man aged 31-35, recently started on treatment for pulmonary tuberculosis, presented with new headache and confusion. CSF showed 20 WBC/µL (77% mononuclear, 23% neutrophils), protein 97 mg/dL, and glucose 42 mg/dL, and CSF GeneXpert MTB/RIF was positive (rifampicin sensitive). Imaging showed hydrocephalus, basilar meningeal enhancement, spinal arachnoiditis, and ischemic infarcts. mNGS detected M. tuberculosis via both shallow and deep sequencing.

## Supplementary Appendix 2: Clinical summaries of CNS infections diagnosed via routine clinical testing that were missed by shallow-depth mNGS

### Patient 049: *Cryptococcus neoformans*

Described in Supplementary Appendix 1.

### Patient 096: *Listeria monocytogenes*

A man aged 46-50 recently diagnosed with multiple myeloma presented with several days of new headache. On admission, CSF analysis demonstrated pleocytosis (105 WBC/µL, 97% mononuclear and 3% neutrophils), elevated protein (97 mg/dL), and glucose 83 mg/dL. MRI brain with and without contrast demonstrated a left temporal abscess with restricted diffusion and surrounding edema. He underwent craniotomy with abscess drainage; subsequent abscess culture grew *Listeria monocytogenes*. This pathogen was missed on both in-country shallow-depth mNGS and deep-sequencing.

## Notes

### Competing Interest Statement

The authors have declared no competing interest.

### Author Declarations

The Institutional review board of University of California San Francisco granted ethical approval for this work. The ethics committee of Hospital Nacional Dos de Mayo granted ethical approval for this work. The ethics committee of Universidad Peruana Cayetano Heredia granted ethical approval for this work.

